# Patient Versus Prediction-Level Evaluation of a Dynamic Clinical Prediction Model of Sepsis

**DOI:** 10.64898/2026.05.26.26354141

**Authors:** Marcelle Tuttle, Carolien C H M Maas, Jennie An, Benjamin S Wessler, William F. Harvey, Harry P Selker, David van Klaveren, David M Kent

**Author notes:** **Corresponding Author:** David Kent.

## Abstract

The Epic Sepsis Model version 2 (ESMv2) is a prediction model embedded into the electronic medical record used to warn clinicians which hospitalized patients are at risk for sepsis. We conducted a retrospective cohort study of 31,951 hospitalizations of 25,760 patients to compare analyses conducted at the commonly used patient-level (where a maximum prediction prior to the onset of sepsis is used to measure performance) vs novel prediction-level (where each prediction is used to measure performance). Sepsis, defined by the Sepsis 3 criteria occurred during 1,049 hospitalizations (3.3%). Patient-level analyses suggested excellent discrimination AUC 0.86; [IQR 0.85, 0.87], whereas prediction-level analyses demonstrated lower performance AUC 0.62; [IQR 0.57, 0.65]. Low estimates of the positive predictive value (14.5% at the patient level vs 4% at the prediction level) imply a high number of false alerts. Common evaluation approaches may overstate the performance of dynamic prediction models and mislead clinical decision-making.

## Introduction

Sepsis is a life-threatening disorder characterized by a dysregulated inflammatory response in response to severe infection resulting in organ dysfunction.^1^ Early recognition of sepsis in hospitalized patients is critical as prompt initiation of fluid resuscitation and antibiotics are associated with reduced mortality.^1,2^ Sepsis guidelines recommend active surveillance of acutely-ill high-risk patients to screen for septic shock.^1^ How best to screen patients, however is an open question.

Recently, machine-learning enabled models imbedded into the electronic medical record (EMR) have been developed to generate dynamic predictions with the goal of proactively warning clinicians about patients who are at risk of sepsis.^3^ The Epic Sepsis Model (ESM) is one such model which when externally validated, demonstrated varying ability to distinguish between low and high-risk patients, expressed as area under the receiver operating characteristics curve (AUC), ranging from worse than random chance to strong discrimination (0.47-0.83).^4–7^ Since these reports, Epic has developed a second model, the Epic Sepsis Model Version 2 (ESMv2) which reportedly demonstrates improved performance over the prior model.

Most studies regarding the performance of these dynamic sepsis prediction models lack a comprehensive approach to their validation. Many authors approach the problem using patient-level validations (e.g., evaluation based on the maximum prediction per hospitalization) as opposed to prediction-level analyses (e.g., evaluation of every predicted score at every time point). These approaches target fundamentally different quantities: patient-level analyses, favored in the clinical literature,^4^ summarizes whether a model ever flags a hospitalization prior to sepsis onset; prediction-level analyses, favored for dynamic prediction in the statistical literature, evaluate whether for a given prediction sepsis occurs within a given time horizon (or time in which sepsis may occur).^8,9^

To date, published external validations of ESMv2 have been limited. One study which included 35,076 emergency department encounters demonstrated that ESMv2 had an AUC of 0.90 for the recognition of sepsis using the patient-level approach.^10^ Another was a prospective validation of 227,091 encounters from 4 hospital systems which demonstrated variable performance of an AUC 0.82-0.92 at the hospitalization level and an AUC 0.75-0.85 at the prediction-level using a 12 hour time horizon.^11^ A validation of the locally-trained version of ESMv2 demonstrated improved performance at both the hospitalization level (AUC 0.89) and prediction-level (AUC 0.84 with an 8 hour time horizon).^12^ Given the paucity of external validations of ESMv2 and the heterogeneity in the analytic approach used in the literature, we sought to validate ESMv2 in a regional hospital system and evaluate the results of patient-level vs prediction-level analyses.

## Methods

### Patient Population

The study sample was derived from the Tufts Medicine system, a network of three hospitals (one tertiary academic and two community hospitals) located in the Boston, Massachusetts metropolitan area. All emergency department and inpatient encounters within the Tufts Medicine system between August 16^th^, 2024 and October 31^st^, 2024 were included in the study sample. Hospitalizations were excluded if the first prediction occurred at the time of onset of sepsis (119 hospitalizations), if there were >2 hours of missing predictions between the time of admission and the first prediction as the timing of the events for these patients was felt to be unreliable (82 hospitalizations). Hospitalizations were also excluded if there was a >1 hour gap in predictions between the last prediction to the onset of sepsis (3 hospitalizations) or discharge (2 hospitalizations). The Tufts Medical Center institutional review board approved the research plan as exempt from review. There was no patient or public involvement in this study.

### Epic Sepsis Model Version 2

The ESMv2 is a gradient boosted tree ensemble model which was derived from 900 candidate variables with 66 features. At 15-minute intervals, a new prediction is generated for each patient with a score on a scale of 0-100 with higher scores denoting higher risk (although not an absolute probability) of developing sepsis. In clinical practice, once the score rises above a certain threshold which is determined by the individual health system, a sepsis alert appears aimed to prompt the clinician to consider initiating therapy for sepsis. At the time of data generation for this study, the ESMv2 was operating in “silent” mode, meaning that it was generating predictions which were not shown to clinical teams. For the purposes of our subgroup analyses and prediction-level validation, a threshold of 25 and above was used to signify that a prediction was positive for sepsis. This threshold was recommended by Epic for the initial evaluation of ESMv2, but health systems ultimately choose individualized threshold scores based on the results of local validation. Additionally, the model can be optimized for a given health system to improve the accuracy of predictions by reweighing the pred included in the model based on the characteristics of each health system’s patients. For the purposes of this study, the “global” or non-localized version of the model was used.

### Sepsis Outcome

Sepsis onset was defined using the Sepsis-3 definition which was defined as the point in time where for each hospitalization a body fluid culture was ordered, antibiotics were ordered, and there was an increase in the Quick Sequential [Sepsis-related] Organ Failure Assessment (qSOFA) score by 2 or more points.^13^ This definition was the same sepsis definition which was used to develop the model.

### Patient-level validation

The patient-level validation was conducted by considering each patient as a unit of analysis. For those patients with the outcome, the highest prediction within the 8-hour window prior to sepsis was validated. For those patients without the outcome, the highest prediction which occurred during the whole hospitalization was used. Model performance was assessed using sensitivity, specificity, positive predictive value (PPV), and negative predictive value (NPV). The number of false alerts to true alerts was ascertained by the equation (1-PPV)/PPV. The discriminative ability of the model, i.e., how well the model can distinguish between low- and high-risk patients, was assessed using AUC. Confidence intervals were obtained using 1,000 bootstrap samples.

Subgroup analyses were conducted by age, sex and race to assess for fairness of the model. Lead time was calculated as the difference from positive alert to sepsis onset in hours.

### Prediction-level validation

While patient-level validations are thought to align with clinical decision making, they may result in inflated AUC statistics as the predictions outside the 8-hour prediction window are discarded. Additionally, as the lengths of stay for patients who do not develop sepsis are often shorter, this may also lead to further inflation of the AUC. Finally, in clinical practice, alerts are often muted for a certain period of time if they are deemed by providers to be false-positives which is difficult to assess in the patient-level validation. To counteract these phenomena, we additionally performed a prediction-level validation where each prediction was individually assessed for its accuracy at predicting sepsis (Supplementary Figure 1). Starting at two hours after admission, the highest prediction within the past two hours was ascertained. The next eight hours were then examined for whether the outcome occurred and statistics for model performance (sensitivity, specificity, PPV, NPV, and AUC) were calculated. This was done for every hour after the two hours following admission. Model performance over time was summarized by a weighted average of the model performance measures, using weights corresponding to the number of observations per hour. Graphs were smoothed using locally weighted polynomial regression. In a sensitivity analysis, model performance was assessed by muting alerts for eight hours after an alert was triggered (i.e., the score threshold of ≥25 was reached). Outcomes were displayed out to 3.5 days as no sepsis events occurred after this time-period.

## Results

There were 31,951 hospitalizations of 25,760 unique patients meeting the inclusion and exclusion criteria (Figure 1). Patients were on average 51.9 (SD 20.9) years old and were predominantly female (54.7%), with White race (62.8%), and Non-Hispanic ethnicity (82.4%) (Table 1).

**Figure 1.**
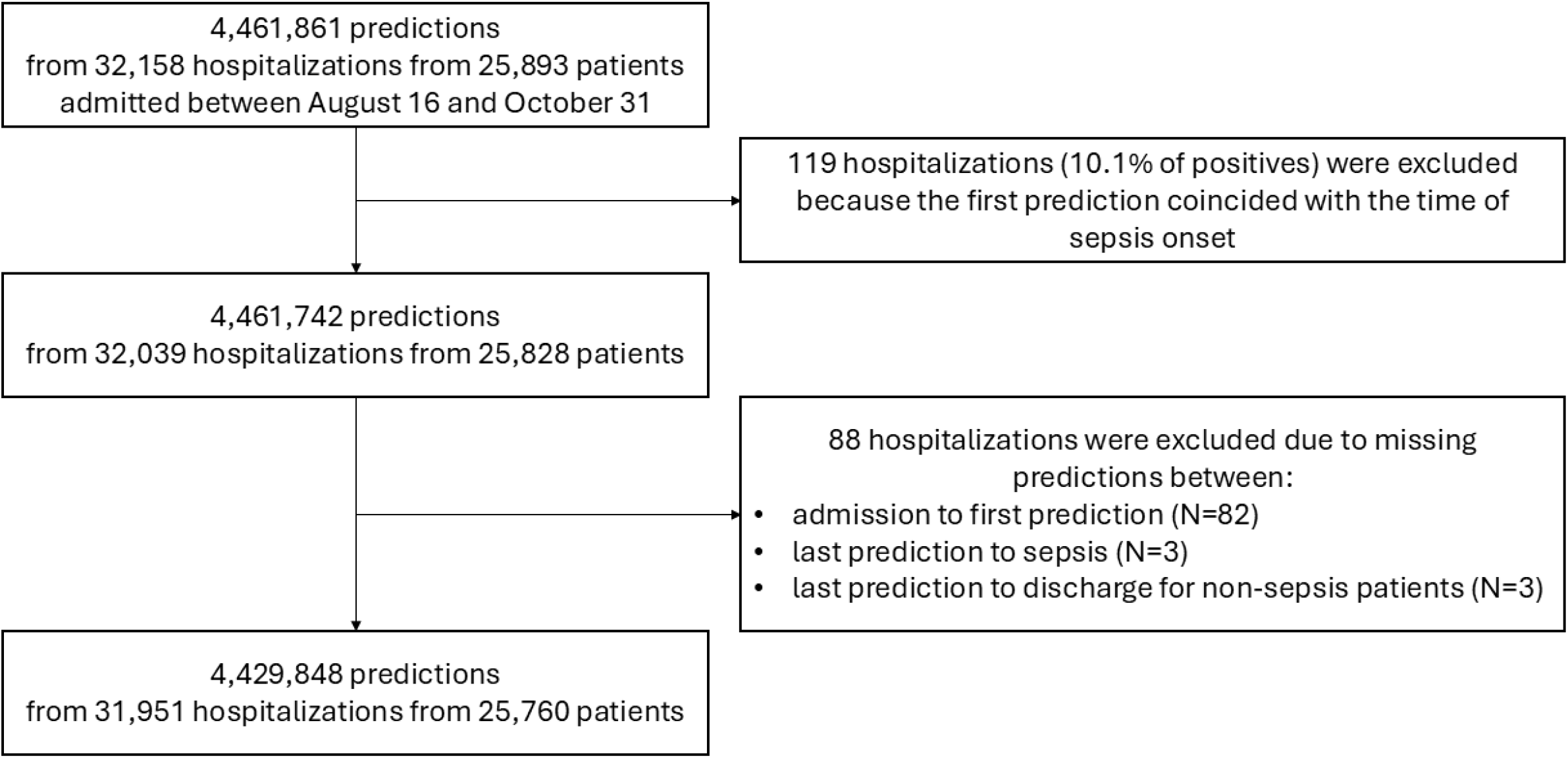
Flow diagram of the study sample. Abbreviations: N = sample size.

**Table 1.** Baseline patient characteristics.

|  | Overall |
| --- | --- |
| Sample size | 26217 |
| Age, mean (SD) | 51.9 (20.9) |
| Females, N (%) | 14329 (54.7) |
| Race, N (%) |  |
| American Indian or Alaska Native | 98 (0.4) |
| Asian | 2738 (10.4) |
| Black or African American | 3190 (12.2) |
| Native Hawaiian / Pacific Islander | 13 (0.0) |
| Other | 3559 (13.6) |
| Unknown | 160 (0.6) |
| White | 16459 (62.8) |
| Patient Ethnicity, N (%) |  |
| Hispanic, Latino/a or Spanish Origin | 4423 (16.9) |
| Not Hispanic, Latino/a, or Spanish Origin | 21595 (82.4) |

Abbreviations: N = sample size; SD = standard deviation.

Sepsis occurred in 1,049 hospitalizations (3.3%) according to the Sepsis-3 definition. These events primarily occurred early during the hospitalizations with 759 (72.4%) occurring within the first 12 hours. Predictions were more likely to be positive early on during the hospitalization, i.e., within the first two hours 13.6% of predictions reached the threshold for an alert. Predictions after the first two hours resulted in alerts 1.7-2.9% of the time. Hospitalizations for patients without sepsis were typically much shorter (i.e., median length of stay 4.7 [IQR 2.7-30.4] hours) as opposed to those with sepsis (i.e., median length of stay 6.8 [IQR 3.8-12.8] days).

When analyzed on the patient-level, most hospitalizations were at low risk for sepsis (i.e., median of the maximum scores per hospitalization was 3, IQR 1-13). As anticipated with a low-frequency outcome, the model had an excellent NPV and a poor PPV (Figure 2). For example, at our chosen threshold of 25, PPV was 14.5% resulting in 6 false alerts for every true alert and the NPV was 98.5% (Table 2). The sensitivity of the model was 62% and the specificity of the model was 87.6%. The AUC was 0.86 [IQR 0.85, 0.87]. For 399 sepsis events (38%), the score exceeded 25 at the time of the event. For 650 sepsis events (62%), the median lead time from the alert firing to sepsis occurring was 2 [IQR 0.75, 5] hours.

**Table 2.**
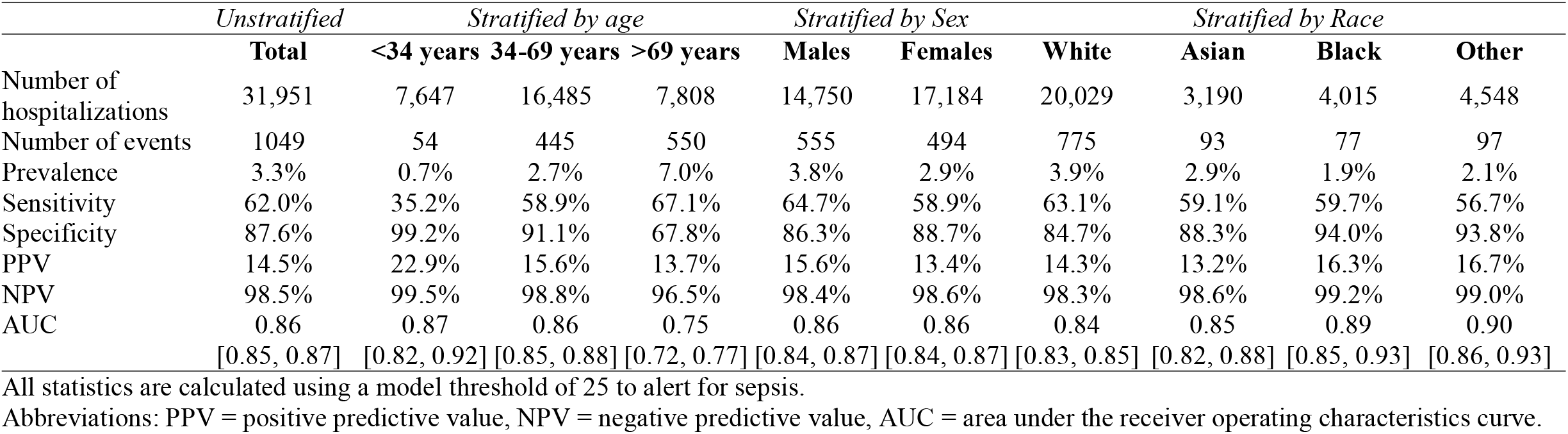
Model performance in subgroups of patients for patient-level validation of ESMv2.

**Figure 2.**
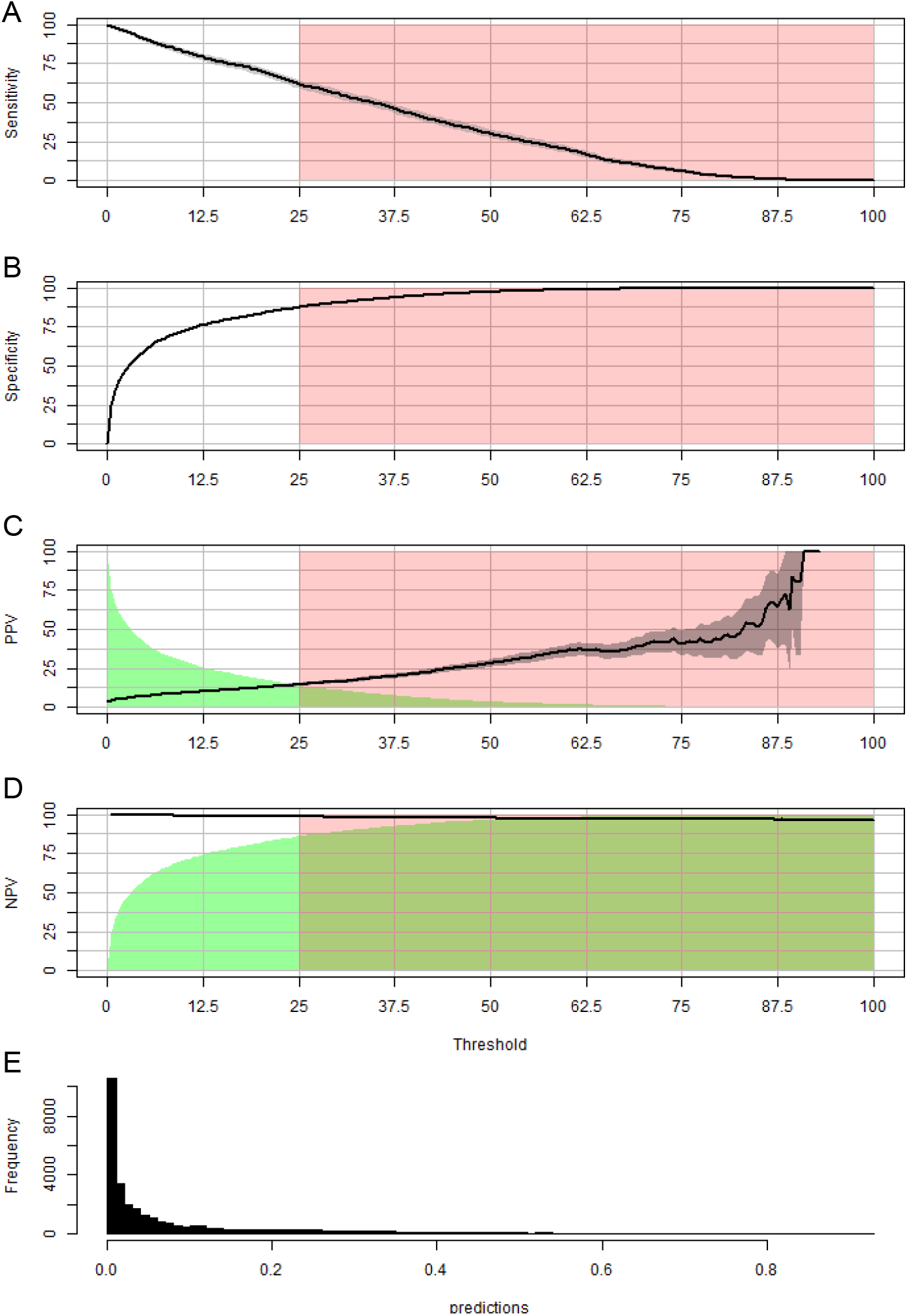
Model performance from the patient-level validation of ESMv2. Graphs depict model performance with A) sensitivity B) specificity, C) PPV, D) NPV and E) the distribution of model predictions. The gray shading around the line represents the 95% confidence interval of each measure. The pink shading denotes the threshold at which the ESMv2 model was deemed to alert for sepsis for the purposes of this study (≥25). The green shading in panels C and D represents the percentage of patients classified as having sepsis or not having sepsis on the PPV and NPV charts respectively. Abbreviations: PPV = positive predictive value, NPV = negative predictive value.

When the study sample was stratified by age, we found that the prevalence of sepsis was much higher in the highest age group vs lowest age group (7.0% vs 0.7%). Additionally, the sensitivity was higher, but the specificity was lower in these patients, with lower PPVs, NPVs and AUCs. Sepsis occurred more frequently in male vs female subjects (3.8% vs 2.9%) although the performance of the model overall was similar. When stratified by race, white patients had the highest prevalence of sepsis (3.9%) and black patients had the lowest prevalence (1.9%). The model performance was similar between race subgroups.

When the performance of the model was gauged using the prediction-level approach, there was overall lower performance with a weighted average AUC of 0.62 [IQR 0.57, 0.65] (Figure 3). Positive predictive values also declined sharply at the very beginning of the hospitalizations resulting in a weighted average of 4%. In other words, there were 24 false alerts for every true alert. The addition of muting alert led to increased instability in the model performance over time and resulted in lower overall performance with a weighted average AUC of 0.53 [IQR 0.52, 0.54] (Supplementary Figure 1).

**Figure 3.**
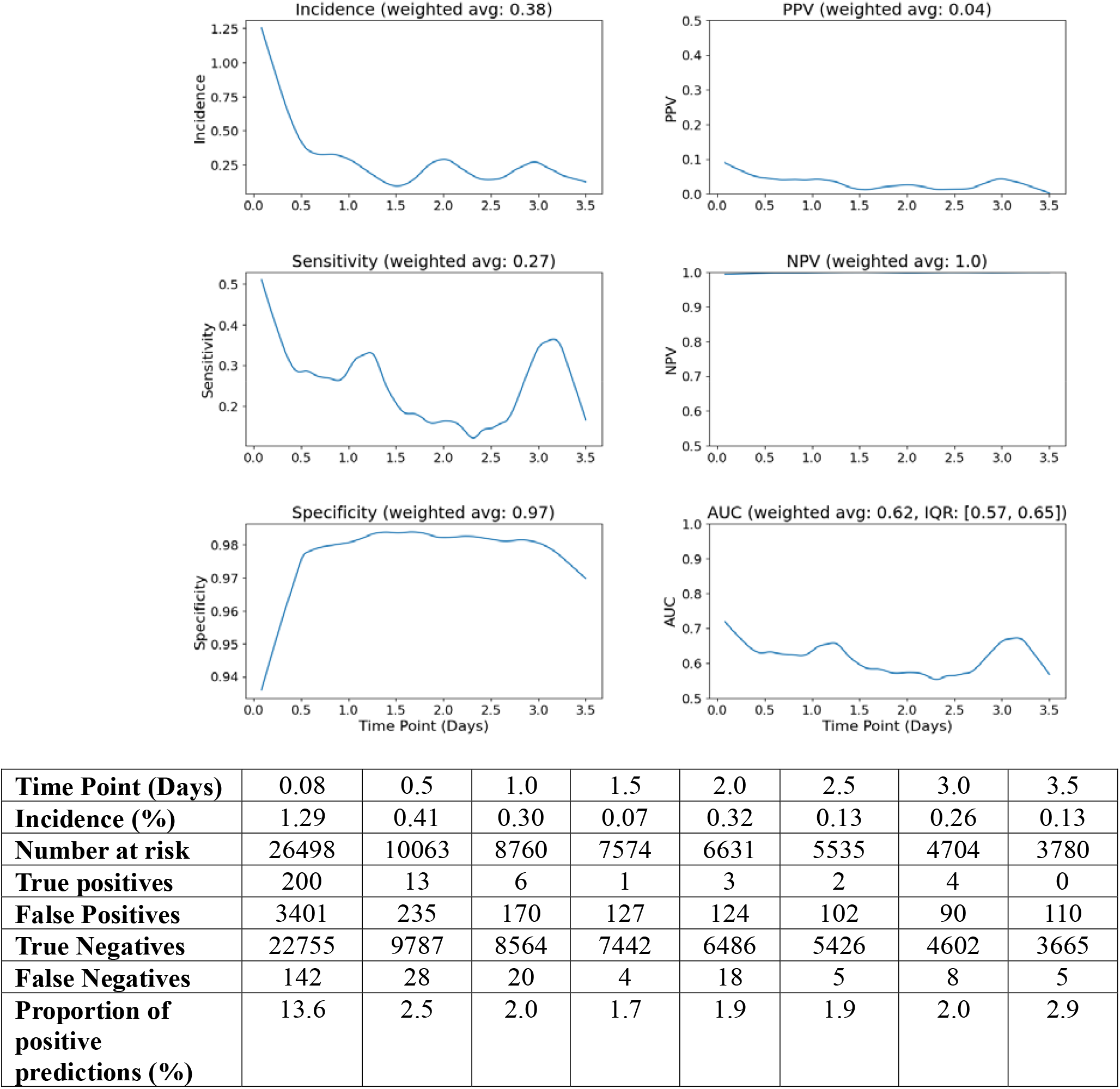
Model performance from the prediction-level validation of ESMv2 without muting the alerts. All statistics are shown using a threshold of 25 to alert for sepsis. Starting at two hours after admission, the highest prediction within the past two hours was ascertained for each hospitalization. Model performance over time was smoothed using locally weighted polynomial regression using a span of 0.15. Abbreviations: PPV = positive predictive value, NPV = negative predictive value, AUC = area under the receiver operating characteristics curve.

## Discussion

When analyzed at the hospitalization level, our external validation of a widely available prediction model for sepsis demonstrated excellent performance with an overall AUC of 0.86. As sepsis is a low-prevalence event in our patient population, however, the PPV for sepsis alerts at a threshold of 25 was also low at 14.5%, indicating most patients flagged as high-risk do not go onto develop sepsis. As sepsis is a low-probability event, it is challenging to develop clinical prediction models with adequate PPV to be clinically useful. When analyzed at the prediction level, we found that the discriminatory capacity of the model was much more modest with a weighted AUC of 0.62. PPV also declined to approximately 4%, corresponding to roughly 24 false alerts for every true alert—levels that would likely be unsustainable in routine clinical care and may contribute to alert fatigue and clinician disengagement.

### Comparison to previous external validations of ESMv2

To our knowledge, despite its widespread adoption into clinical operations, ESMv2 has previously been externally validated in three prior studies. One study of 35,076 emergency department encounters which demonstrated that ESMv2 had an AUC of 0.90, somewhat better performance than in our sample which also included hospitalized in-patients.^10^ This is not wholly unanticipated as there may be differential performance in emergency department encounters vs over the entirety of a patient’s hospitalization as patients seen in the emergency department may be more heterogeneous for sepsis risk compared to those admitted to the hospital. A second study prospectively validated ESMv2 using 227,091 hospitalizations from 4 hospital systems. At the hospitalization level, ESMv2 had variable discriminatory capacity with an AUC of 0.82-0.92, similar to our results.^11^ A third study using the localized version of ESMv2 demonstrated overall improved performance in both the hospitalization level (AUC 0.89) and prediction-level (AUC 0.84 with an 8 hour time horizon) analyses, but is not directly comparable as it has been reweighted to the characteristics of the hospital system in which the analysis occurred.^12^ While prior work has highlighted challenges in evaluating dynamic prediction models using prediction-level approaches,^8,9^ our findings extend this literature in several important ways. First, we demonstrate the magnitude of divergence between commonly used patient-level and novel prediction-level evaluations for a widely deployed clinical model when both evaluations are done on the same population. Second, we show that operational features such as alert suppression can further degrade apparent performance—an issue largely unaddressed in prior work. Third, we link these methodological differences to clinically meaningful consequences, including substantial increases in false-alert burden.

### Patient-level vs prediction-level evaluations

The difference in performance between patient-level and prediction-level analyses arises not only because these approaches estimate different quantities, but also because patient-level evaluation introduces systematic optimistic bias. In particular, patient-level approaches rely on outcome-dependent prediction selection—where the prediction used for evaluation is selected based on future knowledge of when the outcome occurs. This creates a fundamental case–control asymmetry: sensitivity is based on the maximum prediction within a pre-event window for cases, whereas specificity is based on predictions observed over the entire hospitalization for controls. While the sensitivity and specificity obtained in this fashion are interpretable separately, they are not compatible and the resulting AUC does not correspond to any coherent clinical decision problem due to the differences in time horizons. In addition, false positive alerts that occur outside the pre-event window among cases are discarded, and shorter lengths of stay among controls further reduce opportunities for false positives. Together, these mechanisms tend to systematically inflate discrimination metrics. Beyond its effects on model performance, the patient-level approach does not recapitulate the experience that occurs for a clinician who is faced with alerts on the prediction rather than hospitalization level.

While prediction-level approaches avoid outcome-dependent selection and preserve temporal alignment between predictions and outcomes, they are not without limitations. These methods depend on analytic choices such as prediction windows, and time horizons, making them more difficult to interpret in clinical terms. Thus, neither patient-level nor prediction-level evaluation fully resolves the challenges of assessing dynamic prediction models, and different approaches may be appropriate depending on the clinical and operational context.

### Use of ESM in clinical practice

To combat alert fatigue, many hospital systems enable the alerts to be muted for a duration of time. When muting was modeled, there was further reduction in the AUC to 0.53 or essentially no better than random chance. This likely reflects a form of informative suppression: early false-positive alerts, which are common in patients who ultimately develop sepsis, suppress subsequent alerts closer to the period in which the outcome is likely to occur. As a result, patients who go on to develop sepsis may have clinically relevant alerts masked during critical windows for intervention. These findings highlight that model performance is not only a function of the underlying algorithm, but also of how predictions are operationalized within clinical workflows.

### Strengths and Limitations

To our knowledge, our work represents the most comprehensive external validation of ESMv2 applying multiple analytic approaches and scenarios (i.e., muting alerts) to the evaluation of the model. However, our validation is limited to one regional health system and only applies to the global version of the model. It is likely that the performance of the model would improve if it were to be recalibrated to the data within our health system. Second, we also acknowledge that we excluded hospitalizations with missing or irregular prediction streams and these exclusions may be informative, although they make up a small proportion of hospitalizations. Lastly, while we have used a common definition of sepsis which is used broadly for quality improvement work and was used to train this model, sepsis events may have occurred that were not captured by this definition.

## Conclusion

ESMv2 was demonstrated to have variable performance depending on how the data was analyzed in an external validation study. Common evaluation approaches may yield overly optimistic measures of the performance of dynamic prediction models if not interpreted in the context of how predictions are generated and used in practice. While patient-level analyses may remain useful for some purposes, our findings suggest that prediction-level analyses should more routinely complement traditional evaluations of dynamic prediction models, particularly when models are intended to generate repeated clinical alerts over time. Reliance on patient-level metrics alone may overestimate clinical utility and underestimate the operational consequences of false-alert burden.

## Data Availability

All data produced in the present study are available upon reasonable request to the authors

https://github.com/CHMMaas

https://github.com/jennie-TMC/Epic_Sepsis_Validation

## Data availability

Data cannot be made available due to IRB constraints.

## Code availability

The code used to analyze these data are available at: https://github.com/CHMMaas

https://github.com/jennie-TMC/Epic_Sepsis_Validation

## Acknowledgements

MT is funded through the American Kidney Fund Clinical Scientist in Nephrology Fellowship. The project described was supported by the National Center for Advancing Translational Sciences, National Institutes of Health, Award Number UM1TR004398 The content is solely the responsibility of the authors and does not necessarily represent the official views of the NIH.

## Author Contributions

All authors were involved in the conceptualization and design of the study. Analyses were conducted by CM and JA. MT wrote the first draft of the paper with input from all of the authors.

