## Supplemental Material for "Patient Versus Prediction-Level Evaluation of a Dynamic Clinical Prediction Model of Sepsis"

Table of Contents

1. **Supplementary Figure 1**: Diagram of prediction-level analysis

2. **Supplementary Figure 2**. Model performance from the prediction-level validation of ESMv2 with muting alerts for eight hours.

**Supplemental Figure 1**: Diagram of prediction-level analysis

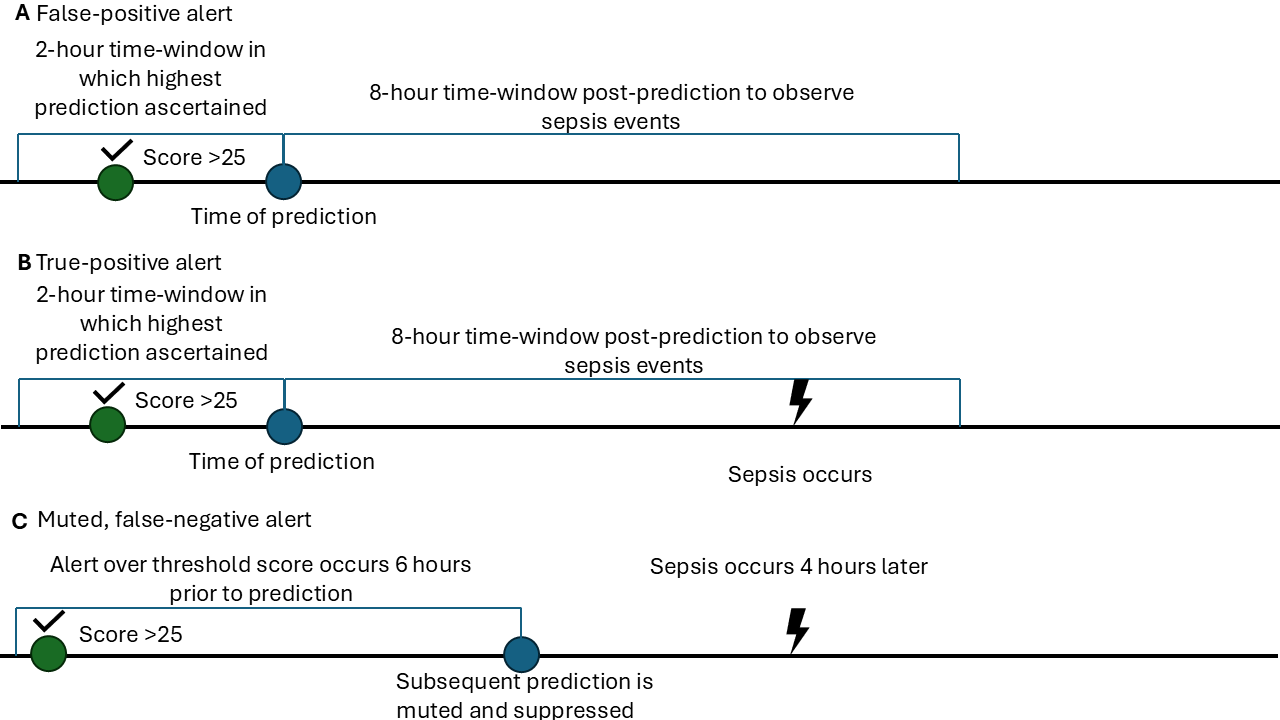

Displayed are schematics of the prediction-level analysis demonstrating A) a false-positive alert, B) a true-positive alert and C) A false-negative alert when muting is modeled. Starting two hours after hospitalization, each prediction serves as its own unit of analysis. The two-hour time window before each prediction is scrutinized for the highest prediction

**Supplementary Figure 2**. Model performance from the prediction-level validation of ESMv2 with muting alerts for eight hours.

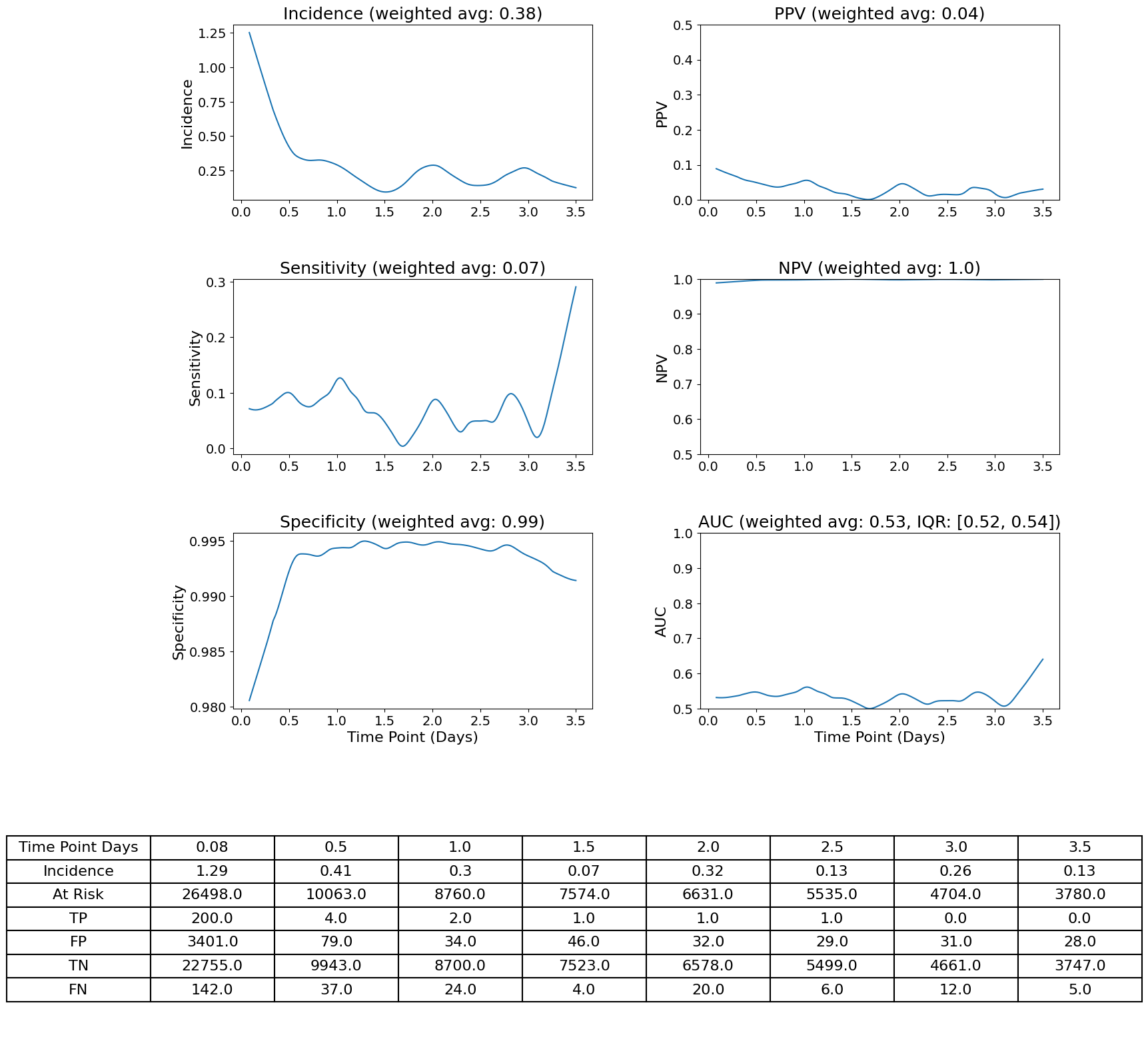

| **Time Point (Days)** | 0.08 | 0.5 | 1.0 | 1.5 | 2.0 | 2.5 | 3.0 | 3.5 |
| --- | --- | --- | --- | --- | --- | --- | --- | --- |
| **Incidence (%)** | 1.29 | 0.41 | 0.30 | 0.07 | 0.32 | 0.13 | 0.26 | 0.13 |
| **Number at risk** | 26498 | 10063 | 8760 | 7574 | 6631 | 5535 | 4704 | 3780 |
| **True positives** | 200 | 4 | 2 | 1 | 1 | 1 | 0 | 0 |
| **False Positives** | 3401 | 79 | 34 | 46 | 32 | 29 | 31 | 28 |
| **True Negatives** | 22755 | 9943 | 8700 | 7523 | 6578 | 5499 | 4661 | 3747 |
| **False Negatives** | 142 | 37 | 24 | 4 | 20 | 6 | 12 | 5 |
| **Proportion of positive predictions (%)** | 13.6 | 0.8 | 0.4 | 0.6 | 0.5 | 0.5 | 0.7 | 0.7 |

All statistics are shown using a threshold of 25 to alert for sepsis. Starting at two hours after admission, the highest prediction within the past two hours was ascertained for each hospitalization. Once an alert was triggered, all predictions for the next eight hours were muted. Model performance over time was smoothed using locally weighted polynomial regression using a span of 0.15.

Abbreviations: PPV = positive predictive value, NPV = negative predictive value, AUC = area under the receiver operating characteristics curve.
